# Efficacy and Viral Dynamics of Tecovirimat in Patients with MPOX: A Multicenter Open-Label, Double-Arm Trial in Japan

**DOI:** 10.1101/2023.08.17.23294241

**Authors:** Yutaro Akiyama, Shinichiro Morioka, Shinya Tsuzuki, Tomoki Yoshikawa, Masaya Yamato, Hideta Nakamura, Masayuki Shimojima, Mizue Takakusaki, Sho Saito, Kozue Takahashi, Mio Sanada, Mika Komatsubara, Kahoru Takebuchi, Etsuko Yamaguchi, Tetsuya Suzuki, Komei Shimokawa, Takeshi Kurosu, Madoka Kawahara, Kohei Oishi, Hideki Ebihara, Norio Ohmagari

## Abstract

**Introduction:** Tecovirimat’s application in treating mpox remains under-researched, leaving gaps in clinical and virological understanding.

**Methods:** The Tecopox study, conducted in Japan, assessed the efficacy and safety of oral tecovirimat therapy in patients with smallpox or mpox. Patients with mpox enrolled between June 28, 2022, and April 30, 2023, were included. We gathered demographic and clinical details along with blood, urine, pharyngeal swab, and skin lesion samples for viral analysis. A multivariable Tobit regression model was employed to identify factors influencing prolonged viral detection.

**Results:** Nineteen patients were allocated to the tecovirimat group. The median age was 38.5 years, and all were male. Ten patients (52.6%) were infected with the human immunodeficiency virus (HIV). Sixteen patients (84.2%) had severe disease. Nine of the 15 patients (60.0%) (four patients withdrew before day 14) had negative PCR results for skin lesion specimens 14 days after inclusion. The mortality rates on days 14 and 30 were 0% and 0%, respectively. No severe adverse events were reported. HIV status and the number of days from symptom onset to tecovirimat administration were associated with lower Ct values (*p* = 0.027 and p < 0.001, respectively). The median number of days when PCR testing did not detect the mpox virus in each patient was 19.5 days.

**Conclusion:** The results suggest that early tecovirimat administration might reduce viral shedding duration, thereby mitigating infection spread. Moreover, patients infected with HIV appeared to have prolonged viral shedding, increasing the transmission risk compared to those without HIV.

**Keypoints:** The Tecopox study revealed that early tecovirimat administration might reduce viral shedding duration, thereby mitigating infection spread. Moreover, patients infected with HIV appeared to have prolonged viral shedding, increasing the transmission risk compared to those without HIV.

## 1. Introduction

As of April 2023, there has been a global reduction in the number of mpox (formerly known as monkeypox) infections. However, the Western Pacific region, especially countries like Japan, South Korea, and China, has witnessed a surge, primarily affecting males [1]. The application of tecovirimat in treating mpox has not been extensively documented in terms of its clinical course or viral load. For instance, among 150 patients with mpox, only two had a history of tecovirimat treatment. Both patients received the medication during hospitalization and were human immunodeficiency virus (HIV)-positive with suppressed viral loads [2].

The duration for which infectious mpox viruses can be detected may vary, with factors such as symptom duration and treatment administration [3]. Nevertheless, there is a scarcity of clinical and virological data regarding the use of tecovirimat for mpox treatment. In a clinical study involving 1,750 patients, 355 individuals were treated with tecovirimat, while three individuals were administered brincidofovir [4]. However, these studies did not assess the viral load in the patients. The absence of viral load evaluation in these studies could limit the assessment of the effectiveness of tecovirimat and brincidofovir as treatments for mpox.

Measurement of viral load is imperative for comprehending its replication and dissemination within the body, as well as for gauging the impact of antiviral medications on suppressing the virus. Further research and clinical studies that incorporate assessments of viral load may offer comprehensive insights into the efficacy of tecovirimat in treating mpox.

Additionally, monitoring viral load levels could prove valuable in determining the duration of viral shedding in patients with varying degrees of disease severity.

## 2. Methods and analysis

### 2.1 Study design and settings

This was a retrospective study involving patients enrolled in the Tecopox study, which is a multicenter, open-label, double-arm trial conducted in Japan (Sapporo, Sendai, Tokyo, Aichi, Osaka, Fukuoka, and Okinawa) that evaluates the efficacy and safety of oral tecovirimat therapy for individuals with smallpox or mpox [5]. Enrollment began with the release of the research database (jRCT, Trial ID: jRCTs031220169) on June 28, 2022, and is scheduled to continue until March 31, 2024. The current study specifically included patients with mpox enrolled in the Tecopox study between June 28, 2022, and April 30, 2023.

### 2.2 Participants

Participants in the Tecopox study were required to meet one to three or all of the following criteria:

1. Obtain written informed consent.
2. Weigh at least 13 kg at the time of providing consent.
3. Have a definitive diagnosis of smallpox or mpox, confirmed by positive polymerase chain reaction (PCR) results from patient specimens.
4. Consent to hospitalization from the start until resolution of the skin lesion.
5. If in the administered group, consent to hospitalization until completion of the 14-day oral tecovirimat therapy.

Patients were excluded from the study if they had any of the following conditions:

1. A history of anaphylaxis to oral tecovirimat or any of its components.
2. Considered unsuitable by the primary investigator.

The Tecopox study consisted of two groups: the Tecovirimat and the Control. Participants voluntarily chose the group they wished to join.

Tecovirimat group: Participants received 14 days of standard supportive care and oral tecovirimat (body weight 13 to <25 kg, 200 mg; 25 to <40 kg, 400 mg; ≥40 kg, 600 mg). Oral tecovirimat was administered within 30 minutes of a moderate-to-high-fat meal.

Control group: Participants received supportive care only.

The Japanese Ministry of Health, Labor and Welfare (MHLW) obtained tecovirimat from a pharmaceutical company, and the study drug was distributed free of charge to each site. If participants wished to switch groups, they could do so by withdrawing their consent and rejoining another group. In such cases, participants in the tecovirimat group stopped taking oral tecovirimat at the time of withdrawal. Since all participants remained in the hospital during the intervention, no specific strategies were employed to improve adherence. All concomitant care and medications were allowed as clinically indicated.

### 2.3 Data collection and measurement

Data were collected on the following patient characteristics: date of birth, age, sex, body weight, race, occupation, medical history, current medications, allergies, history of smallpox vaccination, international travel and sexual contact within the last six months. Samples, including blood, urine, pharyngeal swabs, and skin lesions, were collected for viral examination. For skin lesions, contents from blisters, pustules, and crusts were carefully selected and gathered in accordance with the National Institute of Infectious Diseases (NIID) manual [6]. The most severe skin rash was chosen for sampling. While only one rash was typically selected, multiple samples could be obtained if a patient had several rashes of similar severity, or if a new, more severe lesion emerged during the clinical course. These samples were collected before the first dose of tecovirimat in the treatment group or on the same day of participation in the control group.

Patients were assessed both clinically and daily for adverse events through medical examinations and interviews, which included evaluations of their general condition and any skin lesions. Their general condition was categorized by a researcher as bedridden, weakly ambulatory, or fully ambulatory. Safety evaluations were conducted on days 3, 7, 10, and 14 through blood tests, encompassing white blood cell and differential counts, hemoglobin levels, platelet counts, total bilirubin, aspartate aminotransferase (AST), alanine aminotransferase (ALT), creatinine, and blood sugar levels. For efficacy evaluation, samples of blood, urine, throat swabs, and skin lesions were collected for viral testing. These were repeated on days 21, 30, 60, and 120. Additionally, semen samples were collected for viral testing on days 60 and 120. If a patient found it challenging to visit the hospital, medical examinations and interviews could be substituted with remote methods, such as telephone interviews. In such cases, the collection of physical samples was waived.

Blood testing was conducted at each hospital using equipment standard in clinical settings. All specimens designated for viral testing were stored at temperatures at 2–8 °C after collection and during transport. Alternatively, if immediate shipment was not possible, specimens were stored frozen at temperatures below −20 °C, and preferably below −80 °C, before being transported to the NIID. The copy numbers of the mpox virus were quantified using specific multiplex quantitative PCR (qPCR). The qPCR targeted the F3L gene of the mpox virus, utilizing the following primer and probe sets: forward primer (CATCTATTATAGCATCAGCATCAGA), reverse primer (GATACTCCTCCTCGTTGGTCTAC), and probe (HEX-TGTAGGCCGTGTATCAGCATCCATT-BHQ). Total nucleic acids were extracted from 200 μL samples using a high pure viral nucleic acid kit (Roche Applied Science), following the manufacturer’s protocols, with an elution volume of 50 μL. The qPCR assay involved adding 3 μL of the extracted nucleic acid solution to 23 μL of a reaction mixture containing 2x QuantiTect probe PCR master mix, water provided in the QuantiTect probe PCR kit (Qiagen, Hilden, Germany), and 0.2 μM of each specific primer and probe. The process was performed on a LightCycler 96 (Roche, Basel, Switzerland) with the following conditions: an initial 95 °C for 10 min, followed by 45 cycles of 95 °C for 15 s and 63 °C for 60 s. This method was validated using mpox virus preserved by the NIID.

An independent researcher, free of conflicts of interest, monitored the data. The monitor was chosen from the researchers of the principal investigating institution and had no connection to this study. All adverse events equivalent to CTCAE version 5.0 Grade 3 or higher were recorded, and the relationship between the event and the study was determined.

### 2.4 Primary and secondary endpoint

The primary endpoint of this study is the proportion of patients exhibiting a cycle threshold (Ct) value of 40 or higher in PCR testing of skin lesions at 14 days following enrollment. Secondary endpoints encompass the mortality rate at 14 and 30 days; viral load measurements in blood, throat swabs, skin lesions, and urine at 14, 21, 30, 60, and 120 days; viral load in semen at 60 and 120 days; the duration of fever (≥37.5 °C) starting from the time of enrollment; general condition upon entry, and at 14 and 30 days post-enrollment; and the incidence of adverse events. If more than one skin lesion sample was collected from a participant on the same day, the sample with the lowest Ct value was selected for analysis. Additionally, the study explored factors associated with the Ct values of the skin lesion samples by employing a multivariable regression model.

### 2.5 Statistical analysis

For continuous variables, all patient characteristics as mentioned in section 2.3 were expressed as median and interquartile range (IQR), while categorical variables were represented by absolute values (n) with corresponding percentages (%). We investigated factors associated with prolonged viral detection using a multivariable Tobit regression model. Factors included in this analysis were the HIV status, severity of symptoms due to mpox, the number of days from symptom onset to the day of PCR testing, and the number of days from symptom onset to tecovirimat administration. The dependent variable was the Ct value of skin lesion samples, with the upper limit set at 45. Ct values greater than 45 were classified as “not detected (n.d.)”. In the Tobit model, n.d. values were read as 45 and considered censored. Statistical significance was determined by two-sided p-values less than 0.05. The Tobit model analysis was conducted using R version 4.3.0 [7].

## 3. Results

A total of 27 patients were assessed for eligibility (**Figure 1**). Among these, 19 patients were allocated to the tecovirimat group, and their demographic and clinical characteristics are summarized in **Table 1**. Within this group, 17 patients (89.4%) were Asian, with a median age of 38.5 years, and all were male. Ten patients (52.6%) were infected with the human immunodeficiency virus (HIV), and 13 patients (68.4%) had a history of sexually transmitted diseases. None of the patients had received a smallpox vaccination before the infection. Sixteen patients (84.2%) were categorized as having severe disease, although none had any previously identified risk factors for severe disease. The symptoms upon admission included a rash (94.7%), fever (36.8%), lymphadenopathy (42.1%), malaise (42.1%), severe weakness (38.5%), headache (38.5%), and rectal pain (42.1%). The observed rash sites were the genitals (27.8%), perianal region (44.4%), arms (66.7%), face (66.7%), legs (66.7%), and trunk (77.8%).

**Figure 1.**
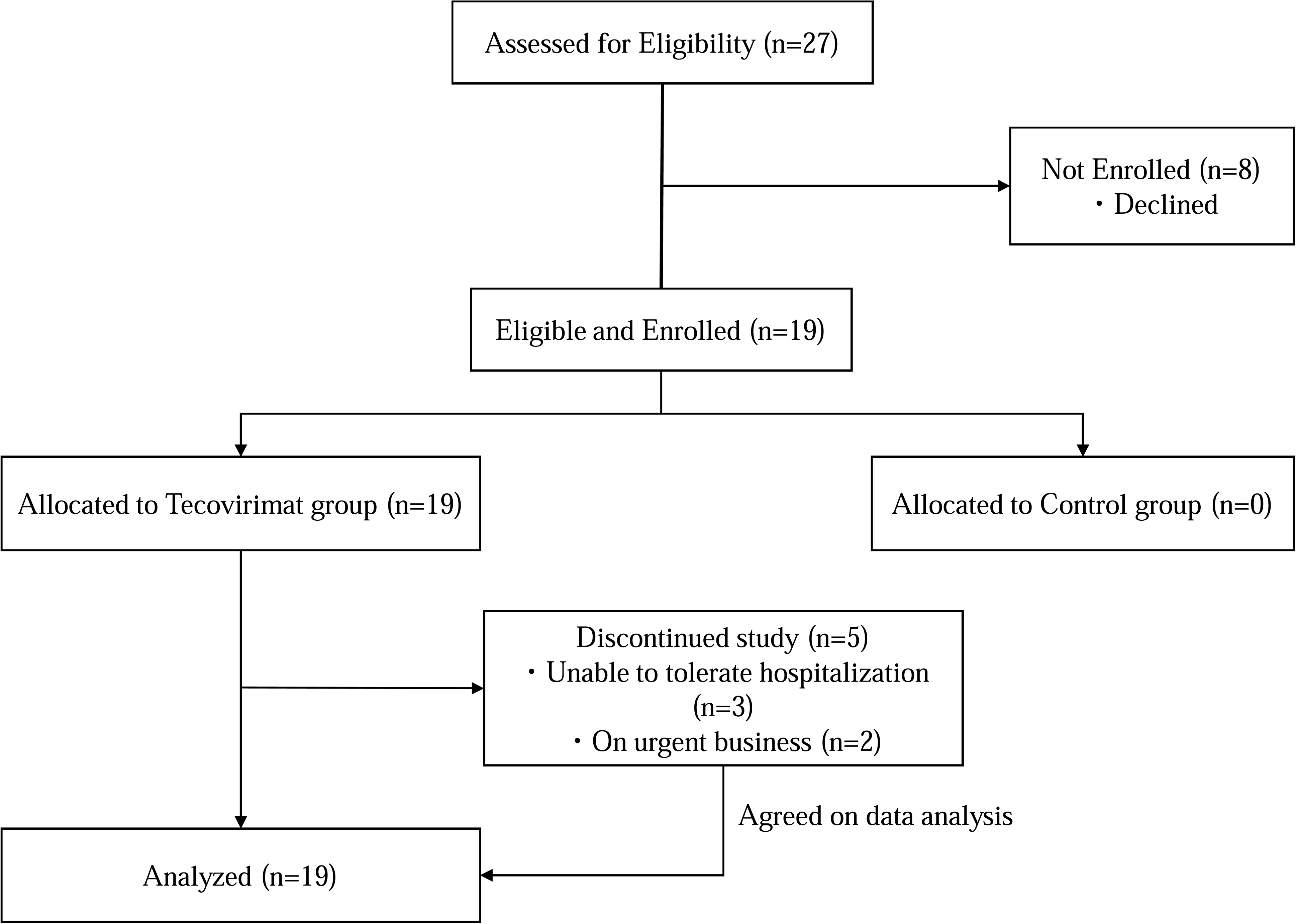
Flowchart of patients’ allocation. Patients opted for eithe Tecovirimat group or control group.

**Table 1.**
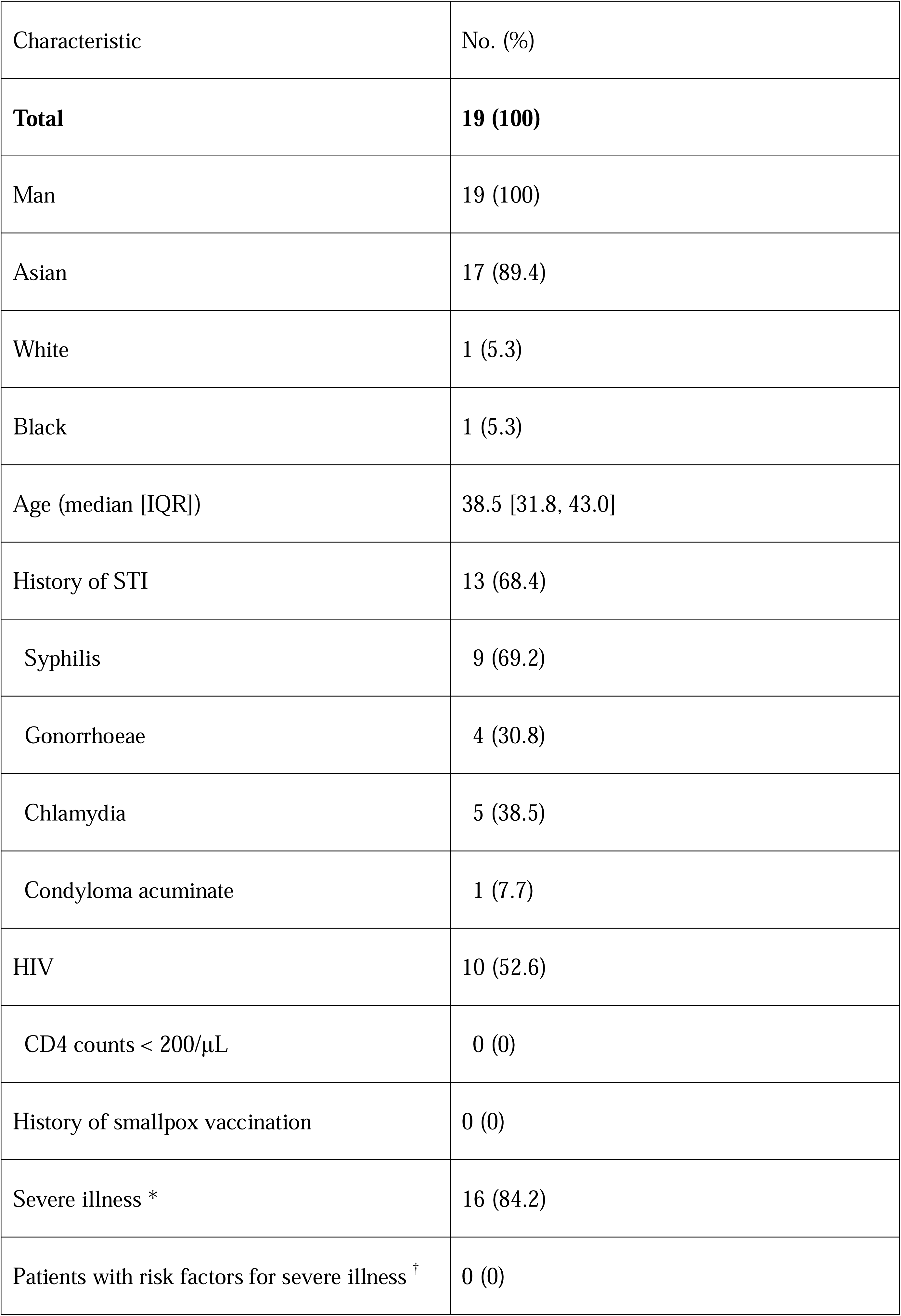
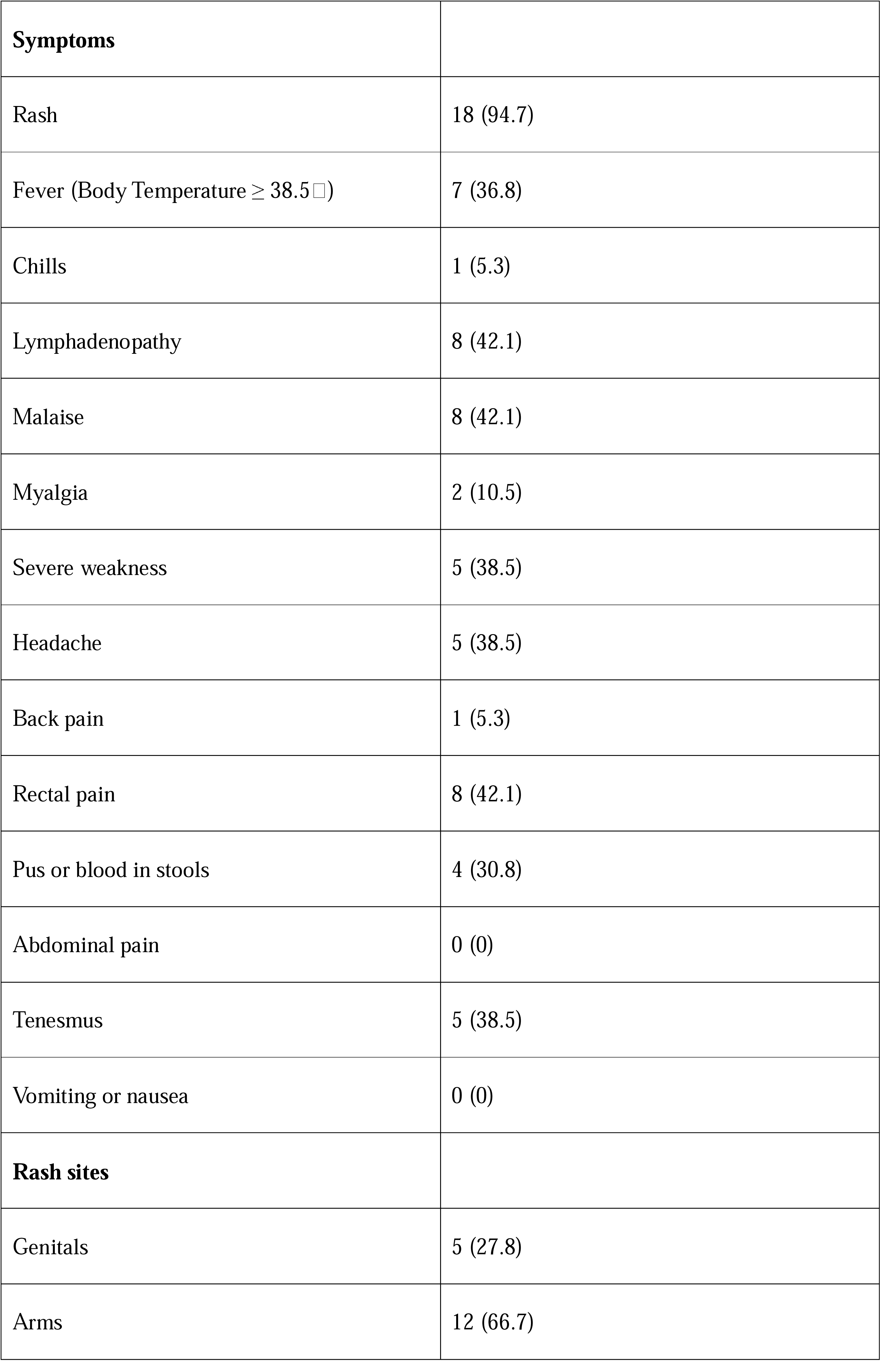
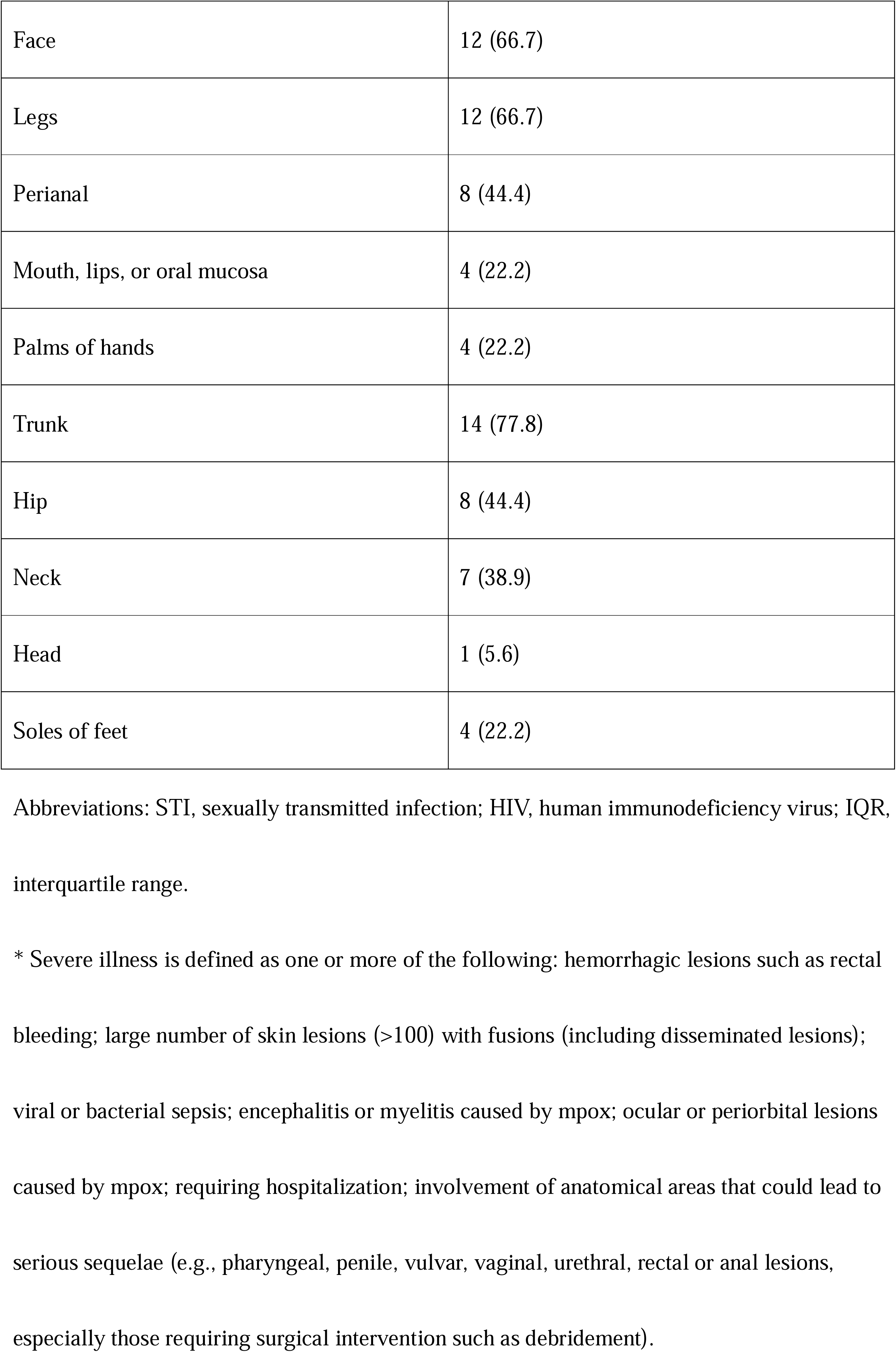
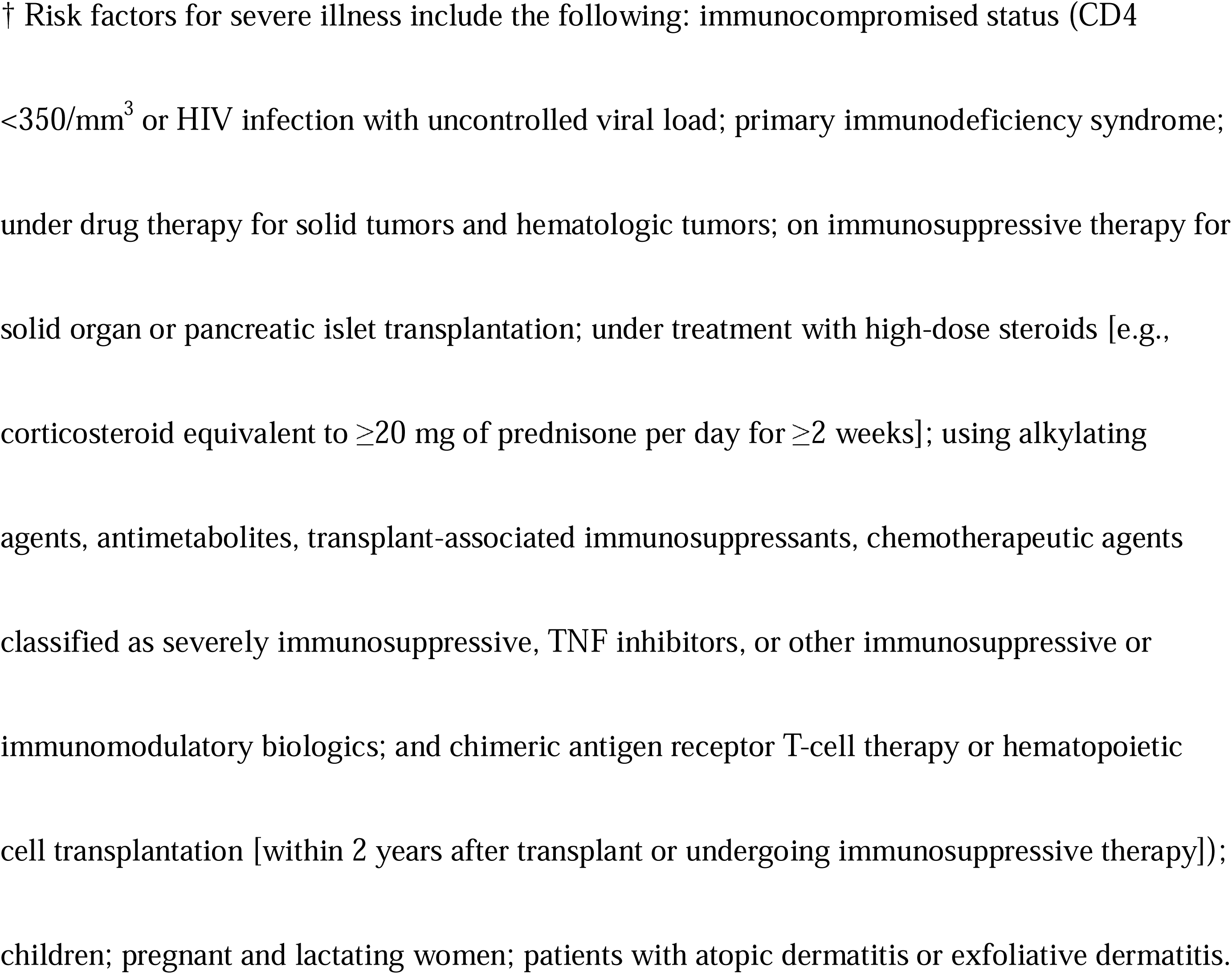
Demographic and clinical characteristics of enrolled patients.

### 3.1. Primary and secondary endpoints

The clinical outcomes for the patients enrolled in the study are summarized in **Table 2**. Out of the participants, five (31.3%) withdrew from the study. With regard to the primary endpoint, nine out of 15 patients (60.0%) (four patients withdrew before day 14) exhibited negative PCR results for skin lesion specimens 14 days post-inclusion. Concerning secondary endpoints, the mortality rates were 0% on both days 14 and 30. No severe adverse events were reported, and only one participant experienced a non-severe adverse event, with none attributed to tecovirimat. The median duration of hospitalization was 14 days, while the median time from rash onset to disappearance of rash was 24 days. The median number of days from symptom onset to viral DNA clearance when PCR testing did not detect the mpox virus in each patient was 19.5 days. The number of samples collected during the Tecopox trial is detailed in **Appendix 1**, and the results of the viral load in the blood, throat swab, skin lesion, and urine at 14, 21, 30, 60, and 120 days, as well as the viral load in semen at 60 and 120 days for each participant, are provided in **Appendix 2**. At the time of study enrollment, mpox virus DNA was detected in 17 cases (89.5%) in skin lesions, 11 cases (57.9%) in throat swabs, 10 cases (52.6%) in blood, and 7 cases (36.8%) in urine. The viral load in the blood, throat swabs, skin lesions, and urine tended to be at its highest upon enrollment, followed by a gradual decrease. No virus was detected in the semen samples on days 60 and 120.

**Table 2.** Clinical outcomes of the enrolled patients.

|  | n (%) |
| --- | --- |
| Withdraw from the study (n=19) | 5 (26.3) |
| <Primary endpoints> |  |
| Negative PCR results (Ct value $\geq 40$) for skin lesion specimens at 14 days after inclusion in the study (n=15) | 9 (60.0) |
| <Secondary endpoints> |  |
| Mortality |  |
| 14 days (n=15) | 0 (0) |
| 30 days (n=14) | 0 (0) |
| Duration of fever ($\geq 37.5^{\circ}\text{C}$) (n=19) | |
| 0 day | 15 (78.9) |
| 1 day | 2 (10.5) |
| 3 days | 1 (5.3) |
| 9 days | 1 (5.3) |
| The general condition at entry (n=19) |  |
| Total ambulation | 19 (100) |
| Partial bedridden | 0 (0) |
| Total bedridden | 0 (0) |
| The general condition at 14 days (n=15) |  |
| Total ambulation | 16 (100) |
| Partial bedridden | 0 (0) |
| Total bedridden | 0 (0) |
| The general condition at 30 days (n=14) |  |
| Total ambulation | 14 (100) |
| Partial bedridden | 0 (0) |
| Total bedridden | 0 (0) |
| Adverse events (n=14) | 0 (0) |
| Duration of hospitalization, days (n=14,<br>median [IQR]) | 14 [14, 15] |
| Time from onset of rash until disappearance<br>of rash, days (n=13, median [IQR]) | 24 [16, 31] |
| Opioids used for analgesia (n=14) | 2 (14.2) |

Abbreviations: PCR, polymerase chain reaction; IQR, interquartile range.

### 3.2. Results of Tobit regression model analysis

HIV status and the time elapsed from symptom onset to tecovirimat administration were correlated with lower Ct values (*p* = 0.027 and <0.001, respectively). Conversely, the number of days from symptom onset to PCR testing was associated with higher Ct values (*p* < 0.001). The details of the analysis are presented in **Table 3**, and the predicted Ct values, corresponding to the interval from the day of symptom onset to the day of PCR testing, are illustrated in **Figure 2**.

**Figure 2.**
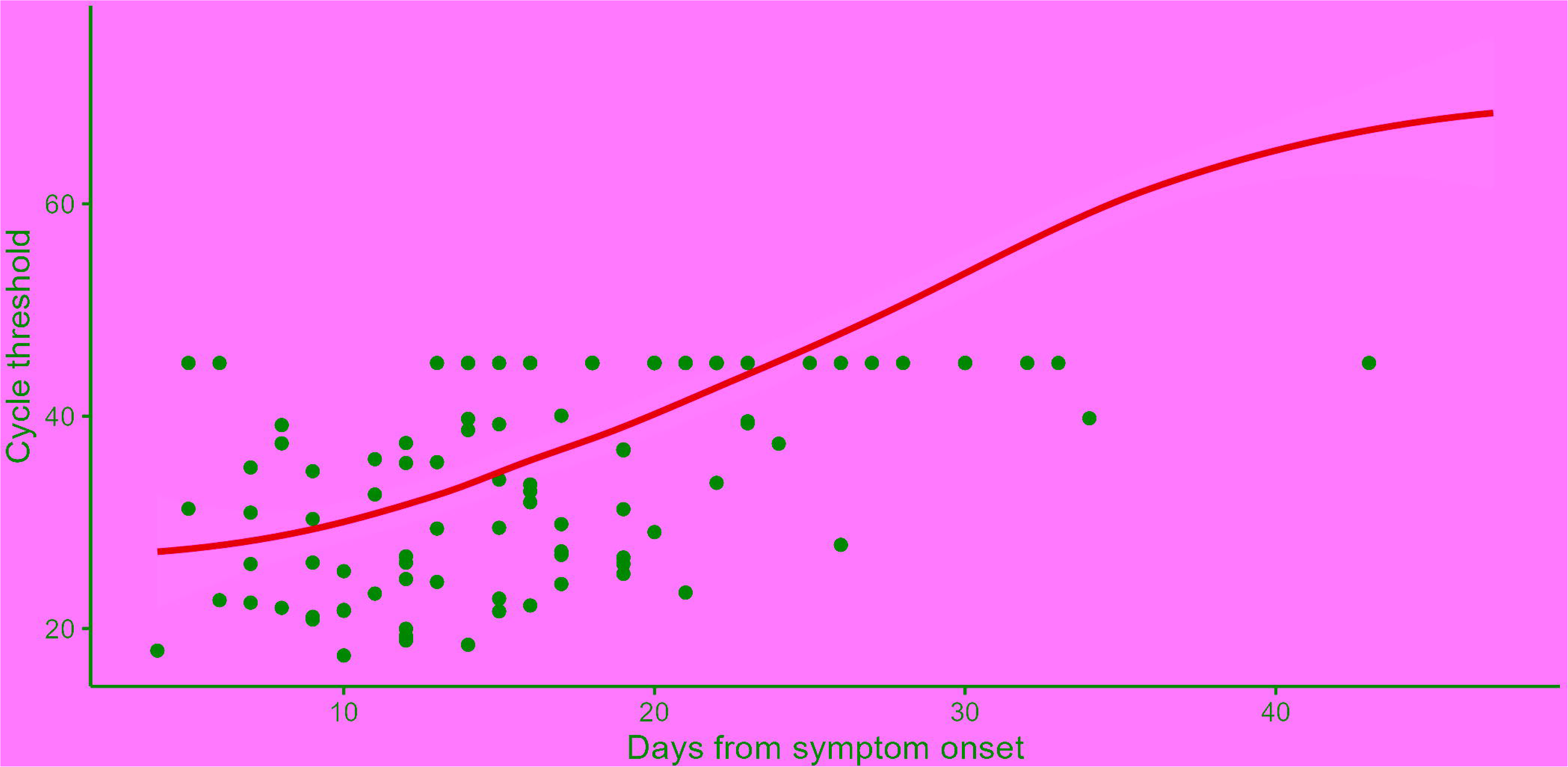
Predicted Ct values of samples from skin lesions according to the interval from the day of symptom onset to the day of PCR testing. The blue line illustrates the predicted Ct values, while the gray areas denote the 95% confidence intervals. The black dots represent the empirical data. Ct; cycle threshold, PCR; polymerase chain reaction

**Table 3.** Results of Tobit regression model analysis.

| Parameter | Coefficient | 95% CI | *P* value |
| --- | --- | --- | --- |
| Positive HIV status | -4.88 | -9.21 to -0.55 | 0.027 |
| Severe symptoms | -5.17 | -11.31 to 0.97 | 0.099 |
| Days from symptom<br>onset | 1.34 | 0.98 to 1.69 | < 0.001 |
| Days from symptom<br>onset to tecovirimat<br>administration | -1.67 | -2.44 to -0.90 | < 0.001 |

Abbreviations: CI, confidence interval; HIV, human immunodeficiency virus

### 3.3. Description of a case with a typical clinical course

In this account, we detail the case of a Japanese man in his 30s who required an extended hospital stay due to a persistent rash. The patient, who had a previous history of syphilis, developed fatigue, fever, and a generalized skin rash after engaging in unprotected sexual activity with men seven days prior to the onset of symptoms. Mpox was diagnosed through a PCR test result from a rash swab. Although his fever subsided two days after diagnosis, the rash spread across his body, accompanied by gradually worsening perianal pain. He was admitted to our hospital five days post-diagnosis and was enrolled in the Tecopox trial. Upon admission, he experienced severe perianal pain and ulcerated lesions. A rectal swab specimen tested positive for *Chlamydia trachomatis* via PCR. Starting from the day of admission (day 0), he was treated with tecovirimat 600 mg twice daily for 14 days for mpox and doxycycline 100 mg twice daily for seven days for chlamydial proctitis. Pain relief for the anal region was provided through the administration of tramadol and non-steroidal anti-inflammatory drugs. On day 14, the patient finished the prescribed course of tecovirimat, and the rash cleared. However, the anal ulcer persisted, preventing discharge. MPOX PCR testing of the anal region, performed on day 19, returned positive results. The patient was finally discharged on day 29, following the resolution of the anal ulcer.

## 4. Discussion

In this study, we detail the clinical progression of 19 patients with mpox and explore viral clearance throughout the illness. We discovered that extended viral detection was correlated with HIV status and the time between symptom onset and tecovirimat administration. This relationship was inversely correlated with the time from symptom onset to the day of PCR testing.

In our dataset, the median days from symptom onset to viral DNA clearance when PCR testing no longer detected the mpox virus in individual patients was 19.5 days (since PCR testing was not conducted daily, the actual day may be slightly earlier). This number is smaller than that found in a previous study [8] but greater than that in another study [9]. Such findings hint that tecovirimat’s effect on the duration of viral shedding remains ambiguous. Nevertheless, our insights from the analysis suggest that administering tecovirimat earlier may reduce the viral shedding duration. While viral DNA detection does not equate to infectivity, it is anticipated to hinder the spread of the infection [5].

In our research, extended viral detection was linked to HIV status. Past reports did not show any difference in treatment results, such as time to improvement or persistent symptom rates, between patients with or without HIV [10]. This suggests that patients with HIV may experience prolonged viral shedding, thereby increasing transmission risk, even though clinical outcomes are consistent between both groups.

In another study, high concentrations of the mpox virus in the blood correlated with a severe clinical course [11]. However, this study revealed that extended viral detection in skin lesions did not correlate with mpox severity, though this relationship might become significant with a larger sample size.

The virus in the semen at 60 and 120 days for eight patients, and at 60 days for one patient, was undetectable. These findings align with the observation that mpox viral DNA was undetectable in all three patients during tecovirimat therapy [12]. The results fortify the evidence that the virus remained undetectable after treatment, suggesting a reduced transmission risk for patients who completed tecovirimat therapy.

Our study had several limitations. Firstly, the absence of randomization in group selection by the participants is a substantial limitation [5], leading to significant allocation imbalance and making an accurate evaluation of tecovirimat therapy’s efficacy challenging. The study’s open-label design may also introduce bias in the efficacy and safety assessment, although we believe the bias was minimal since most primary and secondary outcomes were objectively determined [5]. Secondly, we could not evaluate the clinical outcome of HIV patients with CD4+ counts below 200/µl who underwent tecovirimat therapy. Thirdly, in a previous report, skin lesions appeared at a median of 13 days post-treatment with tecovirimat for mpox. Among six patients tested for orthopoxvirus post-treatment, one tested positive [14]. However, we did not assess post-treatment skin lesions in this study. Therefore, further research is necessary to examine these aspects in greater detail.

## 5. Conclusions

We detail the clinical progression of 19 patients with mpox, investigate the timeline of viral clearance throughout the illness, and identify factors linked to extended viral detection. Our findings suggest that administering tecovirimat earlier may reduce the duration of viral shedding, thereby minimizing infection spread. The study indicates that patients with HIV experienced prolonged viral shedding, leading to a heightened risk of transmission compared to those without infection.

## Acknowledgements

We are grateful to Yusuke Asai for his beneficial suggestions.

## Author contributions

YA, SM, and ST drafted the manuscript. ST performed the statistical analysis. MT, KT, MS, MK, YA, SM, SS, and TS prepared documents for the patients and ensured ethical approval of the study. HE, TY, and MS advised on the sampling methods and viral examinations. NO and SM supervised. All authors participated in the development of the study protocol and approved the final version of the manuscript.

## Ethics approval and consent to participate

This study was reviewed and approved by the Ethics Committee of the Center Hospital of the National Center for Global Health and Medicine (NCGM-C-004505-03). Informed consent was obtained from all participants. All procedures were performed in accordance with the principles of the Declaration of Helsinki.

## Availability of data and materials

All data generated or analyzed during this study are included in this published article and its supplementary 335 information files.

