## Appendix 1 for "Efficacy and Viral Dynamics of Tecovirimat in Patients with MPOX: A Multicenter Open-Label, Double-Arm Trial in Japan"

### Slide 1
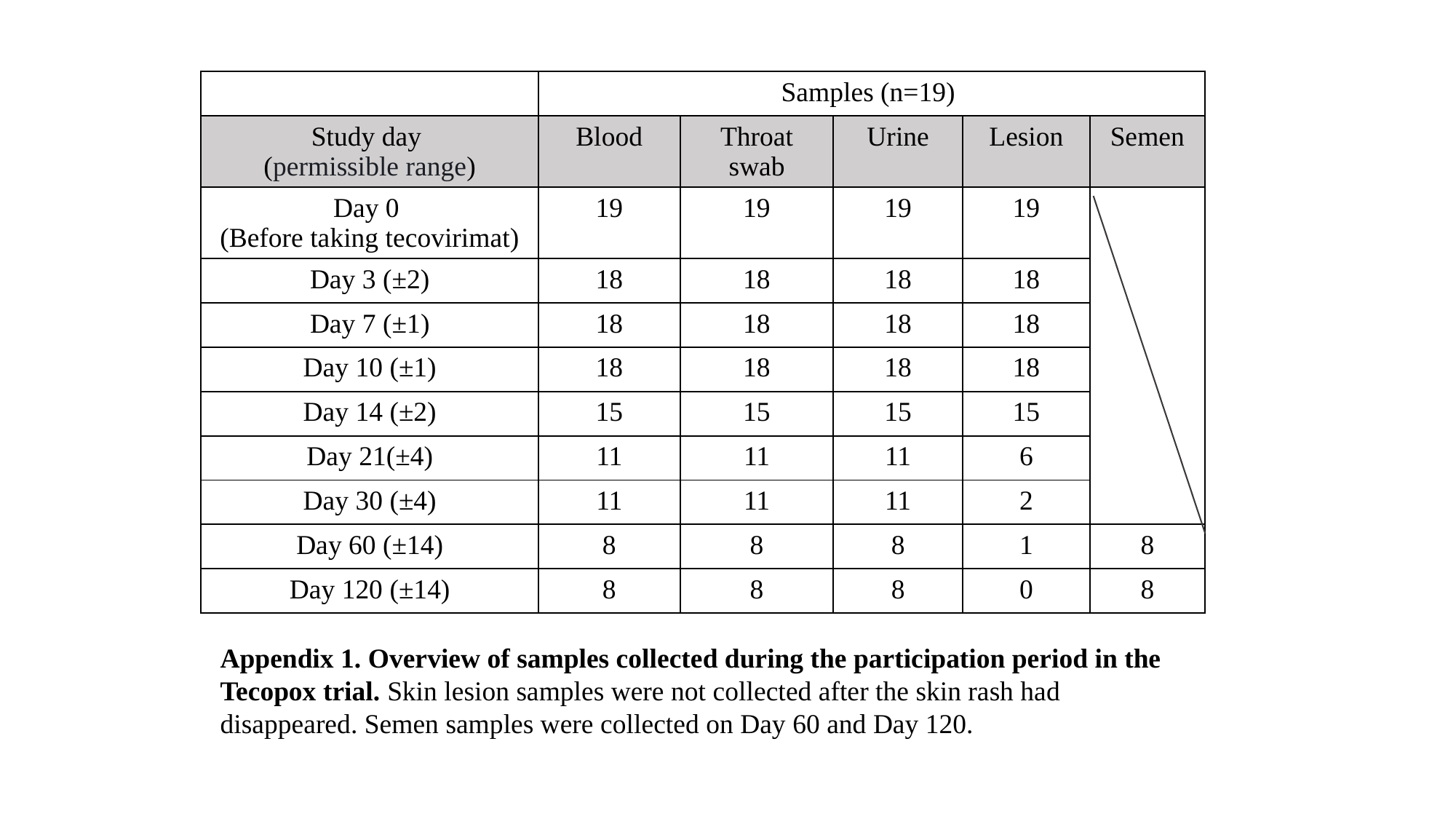

| | Samples (n=19) | | | | |
| --- | --- | --- | --- | --- | --- |
| Study day (permissible range) | Blood | Throat swab | Urine | Lesion | Semen |
| Day 0 (Before taking tecovirimat) | 19 | 19 | 19 | 19 | |
| Day 3 (±2) | 18 | 18 | 18 | 18 | |
| Day 7 (±1) | 18 | 18 | 18 | 18 | |
| Day 10 (±1) | 18 | 18 | 18 | 18 | |
| Day 14 (±2) | 15 | 15 | 15 | 15 | |
| Day 21(±4) | 11 | 11 | 11 | 6 | |
| Day 30 (±4) | 11 | 11 | 11 | 2 | |
| Day 60 (±14) | 8 | 8 | 8 | 1 | 8 |
| Day 120 (±14) | 8 | 8 | 8 | 0 | 8 |
Appendix 1. Overview of samples collected during the participation period in the Tecopox trial. Skin lesion samples were not collected after the skin rash had disappeared. Semen samples were collected on Day 60 and Day 120.
