## Appendix 2 for "Efficacy and Viral Dynamics of Tecovirimat in Patients with MPOX: A Multicenter Open-Label, Double-Arm Trial in Japan"

**Appendix 2. Results of the viral load in blood, throat swab, skin lesion and urine at 14, 21, 30, 60 and 120 days and in semen at 60 and 120 days in each participant**

| **Patients** | ^a^**Days of sample collection** | **Sample type** | | | | |
| --- | --- | --- | --- | --- | --- | --- |
|  |  | **Blood** | **Throat swab** | **Urine** | ^b^**Lesion** | ^c^**Semen** |
| 1 | 0 | negative | 35.15 | negative | 23.28 |  |
|  | 3 | negative | negative | negative | negative |  |
|  | 7 | negative | negative | negative | negative |  |
|  | 10 | negative | negative | negative | negative |  |
|  | 14 | negative | negative | negative | negative |  |
|  | 21 | negative | negative | negative | negative |  |
|  | 30 | negative | negative | negative |  |  |
|  | 60 | negative | negative | negative |  | negative |
|  | 120 | negative | negative | negative |  | negative |
| 2 | 0 | negative | negative | negative | negative |  |
|  | 3 | 40.12 | negative | negative | 34.81 |  |
|  | 7 | negative | negative | negative | negative |  |
|  | 10 | negative | negative | negative | negative |  |
|  | 14 | negative | negative | negative | negative |  |
| 3 | 0 | 37.56 | 31.29 | negative | 19.83 |  |
|  | 3 | negative | negative | negative | 20.40 |  |
|  | 7 | negative | negative | negative | 35.58 |  |
|  | 10 | negative | negative | negative | negative |  |
| 4 | 0 | negative | negative | 39.52 | 21.94 |  |
|  | 3 | negative | negative | negative | 32.61 |  |
|  | 7 | negative | negative | negative | 39.23 |  |
|  | 10 | negative | negative | negative | negative |  |
|  | 14 | negative | negative | negative | negative |  |
|  | 21 | negative | negative | negative |  |  |
|  | 30 | negative | negative | negative |  |  |
|  | 60 | negative | negative | negative |  | negative |
|  | 120 | negative | negative | negative |  | negative |
| 5 | 0 | negative | negative | negative | 29.40 |  |
|  | 3 | negative | negative | negative | 32.92 |  |
|  | 7 | negative | negative | negative | negative |  |
|  | 10 | negative | negative | negative | 39.50 |  |
|  | 14 | negative | negative | negative | negative |  |
|  | 21 | negative | negative | negative | 39.79 |  |
|  | 30 | negative | negative | negative | negative |  |
|  | 60 | negative | negative | negative |  | negative |
|  | 120 | negative | negative | negative |  | negative |
| 6 | 0 | negative | 37.12 | 40.30 | 19.97 |  |
|  | 3 | negative | negative | negative | 22.81 |  |
|  | 7 | negative | negative | negative | 36.86 |  |
|  | 10 | 39.02 | negative | negative | negative |  |
|  | 14 | negative | negative | negative | negative |  |
|  | 21 | negative | negative | negative |  |  |
|  | 30 | 40.27 | negative | negative |  |  |
|  | 60 | 40.72 | negative | 39.66 |  | negative |
|  | 120 | negative | negative | negative |  | negative |
| 7 | 0 | 39.15 | 37.22 | negative | 20.32 |  |
|  | 3 | negative | negative | negative | 19.87 |  |
|  | 7 | negative | negative | negative | 33.24 |  |
|  | 10 | negative | negative | negative | negative |  |
|  | 14 | negative | negative | negative | negative |  |
|  | 21 | negative | negative | negative |  |  |
|  | 30 | negative | negative | 40.00 |  |  |
|  | 60 | negative | negative | negative |  | negative |
|  | 120 | negative | negative | negative |  | negative |
| 8 | 0 | 38.39 | negative | 36.05 | 26.20 |  |
|  | 3 | negative | 39.31 | 31.67 | 19.28 |  |
|  | 7 | negative | negative | 37.91 | 22.16 |  |
|  | 10 | negative | negative | negative | 25.14 |  |
|  | 14 | negative | negative | negative | 39.32 |  |
|  | 21 | negative | negative | negative |  |  |
|  | 30 | negative | negative | negative |  |  |
|  | 60 | negative | negative |  |  | negative |
|  | 120 | negative | negative | negative |  | negative |
| 9 | 0 | 39.93 | negative | 32.55 | 21.74 |  |
|  | 3 | 40.13 | negative | negative | 24.38 |  |
|  | 7 | negative | negative | negative | 27.26 |  |
|  | 10 | negative | negative | negative | 29.07 |  |
|  | 14 | 37.47 | 38.64 | 38.09 | 37.40 |  |
|  | 21 | negative | 36.57 | 38.64 |  |  |
|  | 30 | negative | negative | negative |  |  |
|  | 60 | negative | negative | negative |  | negative |
|  | 120 | negative | negative | negative |  | negative |
| 10 | 0 | 38.70 | 41.52 | negative | 26.07 |  |
|  | 3 | negative | negative | 38.08 | 21.66 |  |
|  | 7 | 37.13 | 36.51 | 35.53 | 38.68 |  |
|  | 10 | 37.46 | negative | 39.89 | 24.18 |  |
| 11 | 0 | 38.59 | 31.32 | 40.23 | negative |  |
|  | 3 | negative | 38.05 | 27.92 | 37.43 |  |
|  | 7 | 37.92 | 36.37 | 36.34 | 26.19 |  |
|  | 10 | 40.83 | 39.43 | negative | 34.01 |  |
|  | 14 | 38.12 | 38.75 | 41.55 | 36.77 |  |
| 12 | 0 | 34.50 | 37.21 | 37.88 | 21.09 |  |
|  | 3 | 36.50 | 37.41 | 37.43 | 37.46 |  |
|  | 7 | negative | negative | 37.94 | 31.88 |  |
|  | 10 | 39.77 | 36.37 | 38.93 | 36.84 |  |
|  | 14 | negative | negative | 36.71 | negative |  |
|  | 21 | negative | negative | negative | negative |  |
|  | 30 | negative | negative | negative |  |  |
|  | 60 | negative | negative | negative |  | negative |
|  | 120 | negative | negative | negative |  | negative |
| 13 | 0 | 33.54 | 38.38 | 37.37 | 29.82 |  |
| 14 | 0 | 40.38 | 30.71 | 38.87 | 24.65 |  |
|  | 3 | negative | 38.55 | negative | 29.48 |  |
|  | 7 | negative | negative | negative | 26.68 |  |
|  | 10 | negative | negative | negative | 33.71 |  |
| 15 | 0 | negative | negative | 24.54 | 20.84 |  |
|  | 3 | negative | negative | 39.94 | 26.79 |  |
|  | 7 | negative | negative | 38.72 | 33.56 |  |
|  | 10 | negative | negative | 41.25 | 31.23 |  |
|  | 14 | negative | negative | negative | negative |  |
|  | 21 | negative | negative | negative | negative |  |
|  | 30 | negative | negative | negative |  |  |
|  | 60 | negative | negative | negative |  |  |
| 16 | 0 | 43.30 | 29.44 | negative | 35.16 |  |
|  | 3 | negative | 36.86 | negative | 25.39 |  |
|  | 7 | negative | negative | negative | 39.74 |  |
|  | 10 | negative | negative | negative | 40.04 |  |
|  | 14 | negative | negative | negative | negative |  |
|  | 21 | negative | negative | negative | negative |  |
|  | 30 | negative | negative | negative |  |  |
| 17 | 0 | 39.60 | negative | negative | 30.90 |  |
|  | 3 | negative | 39.278 | negative | 17.45 |  |
|  | 7 | negative | 39.60 | negative | 18.46 |  |
|  | 10 | negative | negative | negative | 26.92 |  |
|  | 14 | negative | negative | negative | 23.38 |  |
| 18 | 0 | 41.11 | 30.19 | negative | 18.86 |  |
|  | 3 | negative | 36.36 | negative | 21.61 |  |
|  | 7 | 39.42negative | negative | negative | 26.07 |  |
|  | 10 | negative | negative | negative | negative |  |
|  | 14 | negative | negative | negative | 27.87 |  |
|  | 21 | negative | negative | negative | negative |  |
|  | 30 | negative | negative | negative |  |  |
| 19 | 0 | negative | 31.72 | negative | 17.91 |  |
|  | 3 | negative | 39.36 | negative | 22.43 |  |
|  | 7 | negative | 41.66 | negative | 35.94 |  |
|  | 10 | negative | negative | negative | negative |  |
|  | 14 | negative | 39.81 | negative | negative |  |

^a^Days of sample collection since enrollment. Negative: mpox DNA not detected. Values for each specimen: cycle threshold value. Gray mesh: Not available. Samples were not collected after withdrawal from the study.

^b^Skin lesion samples were not obtained after the skin rash disappeared.

^c^Semen samples were collected on days 60 and 120.
